# Strong association between Apgar score at 5 minutes and neonatal survival among at-risk neonates

**DOI:** 10.1101/2020.12.04.20244319

**Authors:** Iván Dueñas-Espín, Andrea Aguilar-Molina, Fernando Aguinaga, Luciana Armijos-Acurio, Ruth Jimbo, Ángela León-Cáceres, María F. Rivadeneira, Silvana Rivera-Guerra, Xavier Sánchez, on behalf of the “Score Bebé” project.

## Abstract

**Objective:** To assess the association between 5-minutes Apgar score and neonatal survival among at-risk neonates.

**Design:** Retrospective survival analysis.

**Setting:** Ecuadorian neonates who died at ≤28 days of life.

**Patients:** We analyzed the nationwide neonatal deaths registered by the Ministry of Public Health of Ecuador between January 2014 to September 2017.

**Main outcome measures:** We performed a survival analysis and estimated adjusted hazard ratios (HR) *per* each 5-minutes Apgar score *stratum*, by Cox proportional hazards models.

**Results:** We included in the study 2893 neonates, 1380 (48%) were female and had a median (P25 to P75) gestational age at birth of 31 (27 to 36) weeks. On univariate analyses, the median survival time in days of life was significantly longer per each increase in the 5-minutes Apgar score, as follows: 0.2 days for ≤4 points, 2 days for 5 points, 2.9 days for 6 points, 3.1 days for 7 points, 3.8 days for 8 points, 4.4 days for 9 points, and 5.5 days for 10 points. On multivariate analyses, and after adjusting for individual and contextual variables, and considering an Apgar score of 9 to 10 points as the reference, the HR was 32% (95% CI: 27% to 37%) higher per each decrease in the Apgar score category of two-to-three points (*p-value* for trend <0.01).

**Conclusions:** There is a strong direct association between Apgar score at 5 minutes and neonatal survival in neonates considered at-risk. This association is independent of gestational age and other neonatal determinants of neonatal mortality.

## Introduction

Scientific evidence shows that the Apgar score is a tool for assessing the response of a newborn during the transition towards extra uterine life. Virginia Apgar, who had the motto of “Nobody, but nobody, is going to stop breathing on me” created the scale in 1953.^1^ The Apgar score comprises 5 components: *(i)* colour; *(ii)* heart rate; *(iii)* reflexes; *(iv)* muscle tone; and *(v)* respiration, and it has been proposed and used as a tool for assessing the neonatal transition, the response to the extra uterine life, and the need for resuscitation.^2^ Specifically, Apgar score helps to add efficacy of immediate resuscitative measures because the score should improve with resuscitation between 1, 5, and 10 or more minutes.^2^

Scientific evidence suggests that the Apgar score is associated with the risk of neonatal and infant death.^3^ Also, a minor degree of physiological dysfunction soon after birth, reflected in the Apgar score, may indicate a slightly higher risk for developmental vulnerability in later childhood.^4^ Additionally, from a public health perspective, this score might serve as a population-level indicator of developmental risk.^5^

Even if the score has been used for decades, considering its possible usefulness for prognosis and follow-up, that has not been completely clarified.^6^ It has been stated that 5-minute Apgar score of 7–10 is reassuring, 4–6 as moderately abnormal, and 0–3 as low score in the term infant and late-preterm infant.^7^ Unfortunately, the accuracy of that categorization in terms of risk stratification has been questioned by several studies.^5,8,9^ Also, an Apgar score of <5 at 5 min appears to be a good predictor of neonatal mortality in infants with a birth weight between 1500 g and 2499 g,^6^ and provide prognostic information about neonatal survival among preterm infants.^10^

Despite gestational age at birth is the most important determinant of neonatal mortality,^11^ Apgar score shows its potential as a tool for estimating prognosis information in non-at-risk^12^ and at-risk neonates.^13^ Under current evidence, we hypothesize that the Apgar score at 5 minutes could be directly and independently associated with survival in at-risk neonates, even considering both: *(i)* the score divided in point-by-point *strata*, and *(ii)* the score divided in two-to-three points categories. The current study aims to assess such association.

## Methods

### Design

We performed a retrospective analysis of all registered deaths of neonates who died at ≤28 days of life and were registered into the Surveillance System of Neonatal Mortality of the Ministry of Public Health of Ecuador, from January 2014 to September 2017. We excluded from the analyses those neonates whose information about their health care attendance and Apgar score at 5 minutes were missed (see the population flow-chart diagram in the **Figure S1** of the **online supplementary material**).

### Population and database

All registered neonatal deaths in public and private settings with available information on individual, contextual, and Apgar score were included in this study. The explanation about the construction of the database is available in the **online supplementary material**.

### Main outcome

We performed a survival analysis by setting the neonatal survival time, as measured by days of life, as the primary outcome. We estimated adjusted hazard ratios (HR) of neonatal death by using: *(i)* Cox’s proportional hazards models and *(ii)* inverse probability weighted (IPW) Cox regression, considering that the analyses were performed on a death registry (*i*.*e*. right truncation).^14^

### Main explanatory variable

Apgar score at 5 minutes was the main explanatory variable. We divided the score in two ways. First, using a point-by-point *strata*, except in the case of the score of ≤4 points which was a single *stratum*. Second, a two-to-three points categories, as follows: *(i)* 0 to 2 points, *(ii)* 3 to 4 points, *(iii)* 5 to 6 points, *(iv)* 7 to 8 points, and *(v)* 9 to 10 points. We did not search for associations between Apgar scores at 1 and 10 minutes and neonatal survival given the lack of information about Apgar score at those time cut-offs.

### Other covariates

Several covariates were available in the database which we classified in two types: *(i)* individual covariates: gestational age at birth, birth weight, small for gestational age (<10th percentile of birth weight for gestational age at birth)^15^ using the Intergrowth equations,^16^ type of delivery, and comorbidities; and, *(ii)* contextual or – the setting’s – covariates: type of health care facility and level of care.

### Statistical analyses and sample size considerations

The analyses were performed using the whole Ecuadorian neonatal mortality data registry from 2014 to 2017; consequently, a sample size calculation was unnecessary. Descriptive statistics were performed using percentages for categorical variables, median and P25 to P75 for discrete variables. To assess the differences of each individual and contextual variables across Apgar score categories, we performed: *(i)* Kruskal Wallis tests for assessing differences of gestational age; *(ii)* ANOVA for assessing differences in birth weight, *(iii)* Chi2 tests for assessing differences of small for gestational age, type of delivery, comorbidities, type of health care center, level of care; and, *(iv)* log-rank test for equality of survivor functions for assessing differences of neonatal survival time, as measured by days.

We estimated crude and adjusted HR with its corresponding 95% confidence intervals (95% CI) *per* each two-to-three points Apgar category.^17^ In that sense, we built: *(i)* multivariate Cox proportional hazards models and *(ii)* inverse probability weighted (IPW) Cox regression [to address the right-truncation (*i*.*e*. death registry)], to evaluate the independent association between Apgar score and neonatal survival time. The model included all the individual and contextual variables, according to the criteria of the researchers.

We performed several secondary analyses to assess the sensitivity of our estimates to our assumptions regarding potential biases, as well as to test for model misspecification. Considering that differential treatment for individual causes of death could affect the estimates, we ran the model excluding neonates who died from *(i)* asphyxia related disorders, *(ii)* congenital malformations, *(iii)* prematurity related disorders, *(iv)* infectious disorders, and *(v)* those who died from other diseases that were not classified in the previous referred groups.

Given the small number of missing data (see footnotes in **Table 1**), we employed a complete case analysis in estimating statistical associations. We considered that there were statistically significant differences when the p-value<0.05. Finally, analyses were performed with Stata 14.2 (Statistical Software Stata: Release 14.2 College Station, TX: StataCorp LP). This study was part of the “Score Bebé” project and conducted with the approval of the Research Ethics Committee in Human Beings (*CEISH*) of the *Pontificia Universidad Católica del Ecuador* (code number: 2018-09-EO).

**Table 1.** Differences in individual and contextual variables of the study population across Apgar scale score at 5 min.

| Variables | Apgar scale score at 5 min<br>(n=2893) |  |  |  |  |  |  | p-value |
| --- | --- | --- | --- | --- | --- | --- | --- | --- |
|  | ≤4<br>(n=763) | 5<br>(n=317) | 6<br>(n=402) | 7<br>(n=417) | 8<br>(n=447) | 9<br>(n=485) | 10<br>(n=62) |  |
| Individual variables<sup>a</sup> |  |  |  |  |  |  |  |  |
| Female sex, n (%)<sup>b</sup> | 392 (51) | 145 (46) | 197 (49) | 197 (47) | 220 (49) | 204 (42) | 25 (40) | 0.05 |
| Gestational age in weeks, median (P25 to P75) <sup>b</sup> | 29 (25 to 34) | 31 (27 to 35) | 30 (27 to 34) | 31 (28 to 36) | 33 (29 to 36) | 35 (31 to 38) | 37 (33 to 39) | <0.01 |
| Birth weight in g, mean (SD) <sup>b</sup> | 1452 (964) | 1580 (914) | 1419 (804) | 1566 (836) | 1738 (800) | 2087 (921) | 2474 (884) | <0.01 |
| Small for gestational age, n (%)<sup>b</sup> | 210 (29) | 76 (25) | 107 (28) | 96 (24) | 105 (25) | 145 (32) | 20 (35) | 0.08 |
| Type of delivery<sup>b</sup> |  |  |  |  |  |  |  |  |
| *C-section, n (%)* | 449 (59) | 178 (58) | 249 (63) | 244 (60) | 274 (62) | 262 (54) | 23 (37) |  |
| *Vaginal delivery, n (%)* | 252 (33) | 110 (36) | 135 (34) | 146 (36) | 147 (33) | 199 (41) | 34 (55) | <0.01 |
| *Dystocic delivery, n (%)* | 56 (7) | 21 (7) | 14 (4) | 19 (5) | 22 (5) | 22 (5) | 5 (8) |  |
| Comorbidities |  |  |  |  |  |  |  |  |
| *Asphyxia related disorders, n (%)* | 222 (29) | 90 (29) | 94 (23) | 105 (25) | 98 (22) | 79 (16) | 7 (11) |  |
| *Malformations, n (%)* | 182 (24) | 74 (23) | 98 (24) | 72 (17) | 106 (24) | 131 (27) | 15 (24) |  |
| *Prematurity related disorders, n (%)* | 272 (36) | 109 (34) | 131 (33) | 147 (35) | 118 (26) | 99 (20) | 7 (11) | <0.01 |
| *Infectious diseases, n (%)* | 63 (8) | 41 (13) | 68 (17) | 78 (19) | 107 (24) | 147 (30) | 29 (47) |  |
| *Other non-previously classified, n (%)* | 24 (3) | 3 (1) | 11 (3) | 15 (4) | 18 (4) | 29 (6) | 4 (6) |  |
| Neonatal survival time in days, P50 (P25 to P75) | 0.2 (<0.1 to 2) | 2 (0.5 to 6.1) | 2.9 (1 to 8) | 3.1 (1. 2 to 8.9) | 3.8 (1.5 to 10.4) | 4.4 (2 to 10.3) | 5.5 (2.5 to 16.1) | <0.01 |
| Contextual variables<sup>a</sup> |  |  |  |  |  |  |  |  |
| Type of health care center |  |  |  |  |  |  |  |  |
| *Private medical care, n (%)* | 287 (38) | 112 (35) | 169 (42) | 111 (27) | 64 (14) | 30 (6) | 0 |  |
| *Public medical care, n (%)* | 476 (62) | 205 (65) | 233 (58) | 306 (73) | 383 (86) | 455 (94) | 62 (100) | <0.01 |
| Level of care<sup>b</sup> |  |  |  |  |  |  |  |  |
| *Primary care unit, n (%)* | 19 (3) | 5 (2) | 5 (1) | 3 (1) | 7 (2) | 12 (2) | 1 (2) |  |
| *Secondary care unit, n (%)* | 289 (38) | 130 (41) | 137 (34) | 184 (45) | 233 (52) | 254 (52) | 44 (71) | <0.01 |
| *Tertiary care unit, n (%)* | 453 (60) | 181 (57) | 257 (64) | 226 (55) | 205 (46) | 218 (45) | 17 (27) |  |

<sup>a</sup> Except for the percentage of neonatal male sex and small for gestational age, there were significant differences of the other (individual and contextual) variables across Apgar scale scores at 5 min, by Chi2, ANOVA, Kruskal Wallis, and Log-rank test for equality of survivor functions, as appropriate. See main text for details. All individual and contextual variables were significantly associated with neonatal survival time in days.
<sup>b</sup> There were missing data (≤5%) in some variables.

## Results

We included in the study 2893 neonates who died between January 2014 to September 2017. Individual and contextual variables of the study population are described in Table **1S**. Of the total study population, 1380 (48%) were female, had a median (P25 to P75) gestational age at birth of 31 (27 to 36) weeks, and had a mean (SD) weight at birth of 1651 (922) g. Regarding their perinatal characteristics, 759 (28%) of them were small for gestational age, most of them (59%) were born by a C-section with a median (P25 to P75) Apgar score at 5 minutes of 6 (4 to 8). The distribution of neonatal deaths per Apgar scale score at 5 minutes is shown in **Figure 1S**. Regarding the comorbidities they developed, prematurity related disorders were most frequent (31%), followed by asphyxia related disorders (24%), congenital malformations (23%), and infectious diseases (18%). Regarding their contextual variables, most deceased neonates were born in public health care facilities (73%) and in secondary and tertiary level hospitals (98%).

**Figure 1.**
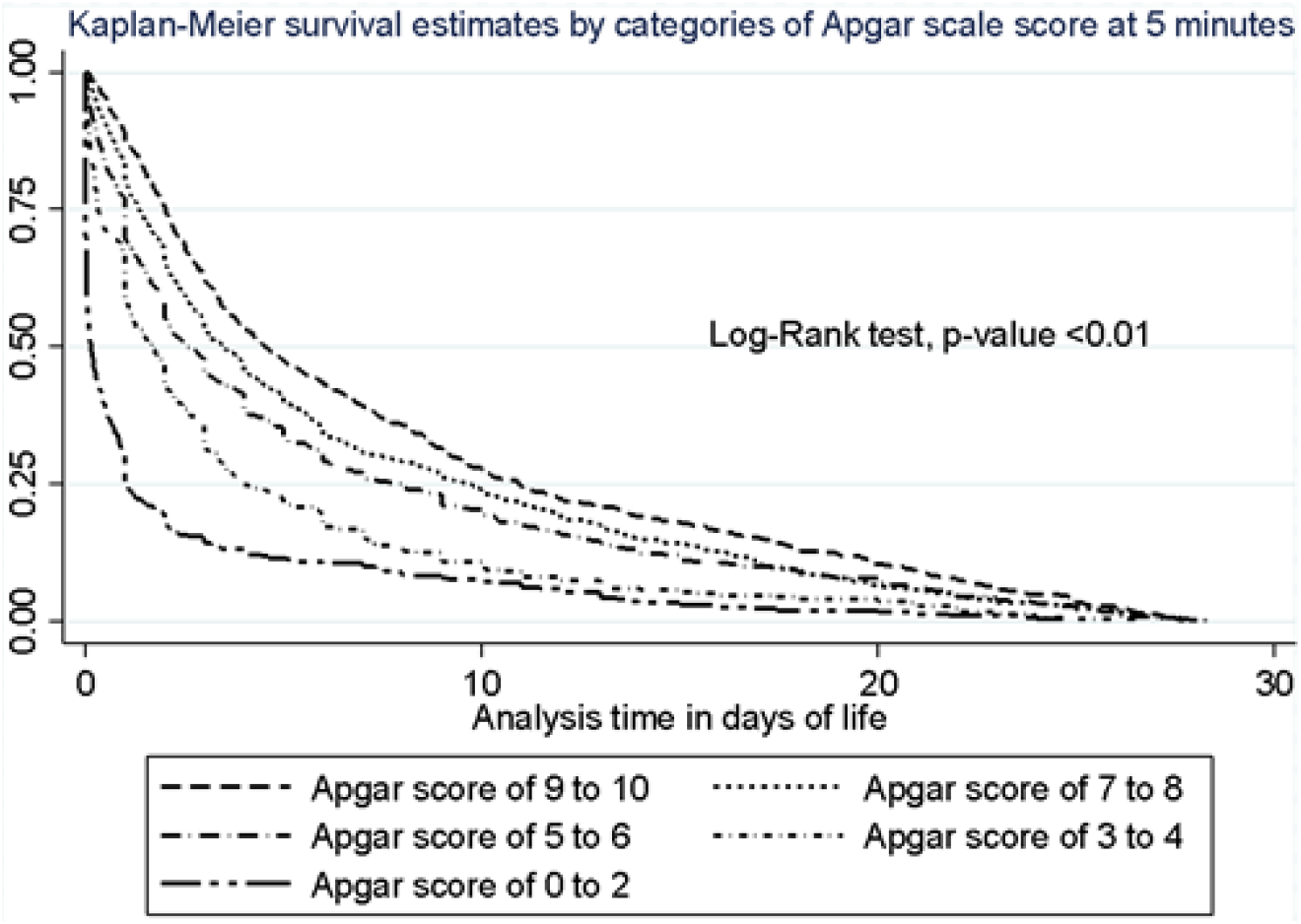
Kaplan Meier survival estimates according to two-to-three points Apgar score at 5 minutes categories.

We found that each explanatory variable was associated with neonatal survival among at-risk neonates during univariate analyses. The median (P25 to P75) survival time in days of life was significantly longer per each increase in the Apgar score a 5 minutes as follows: 0.2 (<0.1 to 2) days when it was ≤ 4 points, 2 (0.5 to 6.1) days when it was 5 points, 2.9 (1 to 8) days when it was 6 points, 3.1 (1. 2 to 8.9) when it was 7 points, 3.8 (1.5 to 10.4) days when it was 8 points, 4.4 (2 to 10.3) days when it was 9 points, and 5.5 (2.5 to 16.1) days when it was 10 points (**Table 1**). Similarly, when using the second categorization of the Apgar score, we found that Kaplan Meier survival estimates were significantly shorter when the Apgar was lower (**Figure 1**).

During multivariate Cox proportional hazards modelling, after adjusting for gestational age, birth weight, small for gestational age, type of delivery, comorbidities and contextual variables, and considering an Apgar score of 9 to 10 points as the reference, the adjusted HR (95% CI lower to upper limits) was 32% (27% to 37%) higher per each decrease in the Apgar score at 5 minutes of two-to-three points category (*p-value* for trend <0.01, see **Table 2**).

**Table 2.**
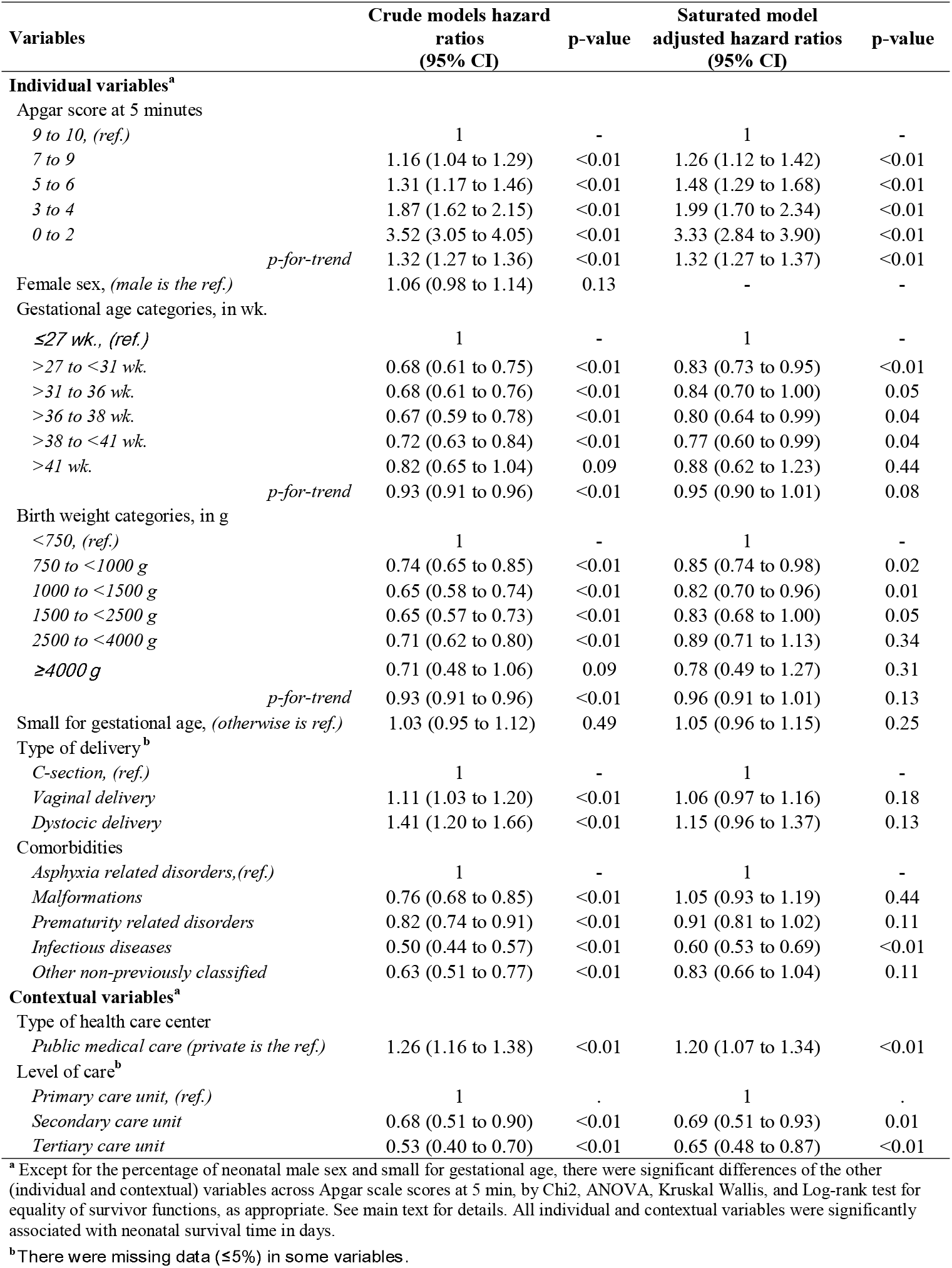
Crude and adjusted hazard ratios according to categories of Apgar scale score at 5 minutes in two to three points categories.

| Variables | Crude models hazard ratios<br>(95% CI) | p-value | Saturated model<br>adjusted hazard ratios<br>(95% CI) | p-value |
| --- | --- | --- | --- | --- |
| **Individual variables<sup>a</sup>** |  |  |  |  |
| Apgar score at 5 minutes |  |  |  |  |
| 9 to 10, (ref.) | 1 | - | 1 | - |
| 7 to 9 | 1.16 (1.04 to 1.29) | <0.01 | 1.26 (1.12 to 1.42) | <0.01 |
| 5 to 6 | 1.31 (1.17 to 1.46) | <0.01 | 1.48 (1.29 to 1.68) | <0.01 |
| 3 to 4 | 1.87 (1.62 to 2.15) | <0.01 | 1.99 (1.70 to 2.34) | <0.01 |
| 0 to 2 | 3.52 (3.05 to 4.05) | <0.01 | 3.33 (2.84 to 3.90) | <0.01 |
| *p-for-trend* | 1.32 (1.27 to 1.36) | <0.01 | 1.32 (1.27 to 1.37) | <0.01 |
| Female sex, (male is the ref.) | 1.06 (0.98 to 1.14) | 0.13 | - | - |
| Gestational age categories, in wk. |  |  |  |  |
| ≤27 wk., (ref.) | 1 | - | 1 | - |
| >27 to <31 wk. | 0.68 (0.61 to 0.75) | <0.01 | 0.83 (0.73 to 0.95) | <0.01 |
| >31 to <36 wk. | 0.68 (0.61 to 0.76) | <0.01 | 0.84 (0.70 to 1.00) | 0.05 |
| >36 to <38 wk. | 0.67 (0.59 to 0.78) | <0.01 | 0.80 (0.64 to 0.99) | 0.04 |
| >38 to <41 wk. | 0.72 (0.63 to 0.84) | <0.01 | 0.77 (0.60 to 0.99) | 0.04 |
| >41 wk. | 0.82 (0.65 to 1.04) | 0.09 | 0.88 (0.62 to 1.23) | 0.44 |
| *p-for-trend* | 0.93 (0.91 to 0.96) | <0.01 | 0.95 (0.90 to 1.01) | 0.08 |
| Birth weight categories, in g |  |  |  |  |
| <750, (ref.) | 1 | - | 1 | - |
| 750 to <1000 g | 0.74 (0.65 to 0.85) | <0.01 | 0.85 (0.74 to 0.98) | 0.02 |
| 1000 to <1500 g | 0.65 (0.58 to 0.74) | <0.01 | 0.82 (0.70 to 0.96) | 0.01 |
| 1500 to <2500 g | 0.65 (0.57 to 0.73) | <0.01 | 0.83 (0.68 to 1.00) | 0.05 |
| 2500 to <4000 g | 0.71 (0.62 to 0.80) | <0.01 | 0.89 (0.71 to 1.13) | 0.34 |
| ≥4000 g | 0.71 (0.48 to 1.06) | 0.09 | 0.78 (0.49 to 1.27) | 0.31 |
| *p-for-trend* | 0.93 (0.91 to 0.96) | <0.01 | 0.96 (0.91 to 1.01) | 0.13 |
| Small for gestational age, (otherwise is ref.) | 1.03 (0.95 to 1.12) | 0.49 | 1.05 (0.96 to 1.15) | 0.25 |
| Type of delivery<sup>b</sup> |  |  |  |  |
| C-section, (ref.) | 1 | - | 1 | - |
| Vaginal delivery | 1.11 (1.03 to 1.20) | <0.01 | 1.06 (0.97 to 1.16) | 0.18 |
| Dystocic delivery | 1.41 (1.20 to 1.66) | <0.01 | 1.15 (0.96 to 1.37) | 0.13 |
| Comorbidities |  |  |  |  |
| Asphyxia related disorders, (ref.) | 1 | - | 1 | - |
| Malformations | 0.76 (0.68 to 0.85) | <0.01 | 1.05 (0.93 to 1.19) | 0.44 |
| Prematurity related disorders | 0.82 (0.74 to 0.91) | <0.01 | 0.91 (0.81 to 1.02) | 0.11 |
| Infectious diseases | 0.50 (0.44 to 0.57) | <0.01 | 0.60 (0.53 to 0.69) | <0.01 |
| Other non-previously classified | 0.63 (0.51 to 0.77) | <0.01 | 0.83 (0.66 to 1.04) | 0.11 |
| **Contextual variables<sup>a</sup>** |  |  |  |  |
| Type of health care center |  |  |  |  |
| Public medical care (private is the ref.) | 1.26 (1.16 to 1.38) | <0.01 | 1.20 (1.07 to 1.34) | <0.01 |
| Level of care<sup>b</sup> |  |  |  |  |
| Primary care unit, (ref.) | 1 | - | 1 | - |
| Secondary care unit | 0.68 (0.51 to 0.90) | <0.01 | 0.69 (0.51 to 0.93) | 0.01 |
| Tertiary care unit | 0.53 (0.40 to 0.70) | <0.01 | 0.65 (0.48 to 0.87) | <0.01 |

<sup>a</sup> Except for the percentage of neonatal male sex and small for gestational age, there were significant differences of the other (individual and contextual) variables across Apgar scale scores at 5 min, by Chi2, ANOVA, Kruskal Wallis, and Log-rank test for equality of survivor functions, as appropriate. See main text for details. All individual and contextual variables were significantly associated with neonatal survival time in days.
<sup>b</sup> There were missing data (≤5%) in some variables.

A similar increase in the HR *per* each decrease in the Apgar category was found when we stratified the analysis across gestational age at birth categories, as follows: 41% (34% to 48%, *p-value* for trend <0.01) increase when neonates had <34 weeks of gestational age, 39% (25% to 54%, *p-value* for trend <0.01) increase when neonates had 34 to <37 weeks, 24% (14% to 34%, *p-value* for trend <0.01) increase when neonates had 37 to <41 weeks. For those who had ≥41 weeks (n=56) the hazard ratio was higher in most strata by comparison with a better Apgar score (9 to 10) at 5 minutes (**Table 3** and **Figure 3S)**, but the trend was not statistically significant (*p-value* for trend = 0.97).

**Table 3.** Adjusted hazard ratios per categories of Apgar scale score at 5 minutes, stratified by gestational age at birth.

| Apgar scale score<br>at 5 minutes | Adjusted hazard ratios (95% CI) per gestational age categories<sup>a</sup> |  |  |  |  |  |  |  |
| --- | --- | --- | --- | --- | --- | --- | --- | --- |
|  | <34 wk.<br>n=1790 | p-value | 34 to <37 wk.<br>n=412 | p-value | 37 to <41 wk.<br>n=625 | p-value | ≥41 wk.<br>n=56 | p-value |
| 9 to 10, (ref.) | 1 | - | 1 | - | 1 | - | 1 | - |
| 7 to 9 | 1.27 (1.07 to 1.52) | <0.01 | 1.26 (0.95 to 1.67) | 0.11 | 1.36 (1.08 to 1.71) | <0.01 | 1.68 (0.50 to 5.66) | 0.40 |
| 5 to 6 | 1.51 (1.26 to 1.83) | <0.01 | 1.55 (1.10 to 2.19) | 0.01 | 1.74 (1.33 to 2.28) | <0.01 | 3.02 (0.46 to 19.85) | 0.25 |
| 3 to 4 | 2.35 (1.88 to 2.96) | <0.01 | 1.82 (1.22 to 2.71) | <0.01 | 1.77 (1.25 to 2.51) | <0.01 | 5.86 (1.18 to 29.13) | 0.03 |
| 0 to 2 | 3.93 (3.17 to 4.88) | <0.01 | 8.11 (4.92 to 13.35) | <0.01 | 2.34 (1.64 to 3.35) | <0.01 | 0.74 (0.11 to 5.06) | 0.75 |
| *p-for-trend* | 1.41 (1.34 to 1.48) | <0.01 | 1.39 (1.25 to 1.54) | <0.01 | 1.24 (1.14 to 1.34) | <0.01 | 0.99 (0.69 to 1.43) | 0.97 |

<sup>a</sup> Each column is a different model adjusted by variables of Table 2: individual variables: (i) gestational age, (ii) birth weight, small for gestational age, (iii) type of delivery, (iv) comorbidities; and contextual variables: (i) type of health care center, (ii) public or private care, (iii) level of care. Please see main text for details.

When we performed the IPW Cox proportional hazards models after artificial cutting-off of the follow up at 21 days, the adjusted survival estimates yielded very similar results corroborating a “dose-response” shape in the association between Apgar score and survival time in days of life, at both, the whole sample and stratified by gestational age at birth categories (**Table 4** and **Figure 4S**).

**Table 4.** Adjusted estimates of survival, in days of life, across gestational age categories by IPW Cox proportional hazards models after artificial cutting-off the follow up at 21 days.

| Apgar scale score | Estimated survival in days (95% CI) of life according to IPW Cox Proportional Hazards models<sup>a</sup> |  |  |  |  |  |  |  |
| --- | --- | --- | --- | --- | --- | --- | --- | --- |
|  | Whole sample<br>n=2893 | p-value | <34 wk.<br>n=1790 | p-value | 34 to <37 wk.<br>n=412 | p-value | 37 to <41 wk.<br>n=625 | p-value |
| 9 to 10 | 5.36 (4.72 to 6.00) | <0.01 | 5.23 (4.26 to 6.19) | <0.01 | 6.67 (5.38 to 7.95) | <0.01 | 6.35 (5.17 to 7.53) | <0.01 |
| 7 to 9 vs. 9 to 10 | -0.27 (-1.01 to 0.46) | 0.46 | 0.31 (-0.78 to 1.40) | 0.57 | -1.69 (-3.25 to -0.13) | 0.03 | -1.95 (-3.33 to -0.56) | <0.01 |
| 5 to 6 vs. 9 to 10 | -1.26 (-2.03 to -0.49) | <0.01 | -1.15 (-2.21 to -0.09) | 0.03 | -2.05 (-3.97 to -0.13) | 0.03 | -2.67 (-4.38 to -0.96) | <0.01 |
| 3 to 4 vs. 9 to 10 | -2.58 (-3.37 to -1.80) | <0.01 | -2.18 (-3.37 to -0.99) | <0.01 | -4.63 (-6.22 to -3.03) | <0.01 | -3.73 (-5.19 to -2.27) | <0.01 |
| 0 to 2 vs. 9 to 10 | -2.82 (-3.79 to -1.85) | <0.01 | -2.39 (-3.74 to -1.04) | <0.01 | -5.96 (-7.28 to -4.63) | <0.01 | -1.88 (-5.21 to 1.45) | 0.26 |

<sup>a</sup> Each column is a different IPW Cox model, adjusted by variables of Table 2: individual variables: (i) gestational age, (ii) birth weight, small for gestational age, (iii) type of delivery, (iv) comorbidities; and contextual variables: (i) type of health care center, (ii) public or private care, (iii) level of care. Please see main text for details.

Sensitivity analyses performed to assess if the estimates changed with specific exclusions yielded similar results too (**Table 2S**). Consequently, excluding *(i)* asphyxia related disorders, *(ii)* congenital malformations, *(iii)* prematurity related disorders, *(iv)* infectious disorders, and *(v)* other disorders non previously classified, did not change the estimates.

## Discussion

The Apgar score has a strong potential for neonatal mortality prognosis among at-risk neonates because it was directly associated with neonatal survival. Scientific evidence suggests that it is associated with long-term neurodevelopment outcomes^5^ and long-term mortality.^3^ In that sense, we support its usefulness for survival prognosis among at-risk neonates, even when using more granular categorizations of the Apgar score, like point-by-point and two-to-three points strata. Also, we demonstrated that this association is independent of gestational age at birth and other individual and contextual determinants.

Our results show similarities with other studies about Apgar prognosis in at-risk neonates; for example, it has been previously demonstrated that premature neonates with a low 5-minute Apgar score have higher risk of mortality and of receiving intensive care.^6,18^ Other studies demonstrated association between the Apgar score and mortality in neonates, infants, and late childhood.^3^

There is no standardized categorization of the Apgar score at 5 minutes. However, our findings suggest that a more disaggregated categorization would help to estimate complications and death among at-risk neonates; and probably, among infants and children.

It is important to recognize that there is much left to be done to reduce preventable neonatal death in Latin American and the Caribbean,^19^ like substantially improving access to health care services, assuring universal facility-based delivery, and enhancing emergency obstetric and timely neonatal interventions.^20^ In the specific case of Ecuador, the lack of such effective interventions have affected neonatal mortality causing its increase from 4.6 to 6.0 neonatal deaths *per* 1000 live newborns since 2014 to 2019.^21^ In that sense, enhancing newborn care is an outstanding debt from the State to society, and urgent interventions are required.

Also, Apgar score assessment, and particularly a change in the score in between 1 minute and 5 minutes, is a useful index of the response to resuscitation.^2^ A standardized Apgar assessment for helping decision-making processes during the early care of at-risk neonates should assure timely health care interventions across Apgar bands of risk. Therefore, it is possible that Apgar and neonatal care training for health care workers, appropriate supply of inputs, and a substantial improve in communication during neonatal transport, could substantially contribute to reduce the neonatal mortality rate in Ecuador.

Our study had several strengths. Using a nationwide database assures statistical power and a proper inclusion of the whole casuistic of at-risk neonates in Ecuador. Also, our study included several individual and contextual covariates as potential confounders, reducing the risk of confounding bias.

As a main limitation, we recognize that it is difficult to assess the quality of the Apgar score information that we used in our analyses. It is important to mention that, despite the lack of specific knowledge about the training on the Apgar score assessment of health care workers, the current Ecuadorian regulations include Apgar assessment as a compulsory process.^22,23^ This has been used in the country for more than 25 years. Therefore, it is a common and very interiorized process for most health care professionals at different levels of care and in private and public settings.

Another limitation is the lack of information about Apgar score at 1 and 10 minutes of life, which precludes a more comprehensive assessment of Apgar as a prognosis tool. Even if a potential source of selection bias is possible due to the use of a death registry database, we employed the IPW Cox proportional hazards modelling which resulted in similar estimates than our fixed-effects Cox models. Consequently, we consider that right-truncation is a non-significant source of bias.

Finally, the lack of non-at-risk neonates in the analyses precludes a proper assessment of the usefulness of score in healthier neonates. Nonetheless, previous evidence^3,12^ supports our findings, indicating that Apgar score could be used as a prognosis tool of neonatal mortality at both, at-risk and non-at-risk neonates.

## Conclusion

There is a strong direct association between Apgar score at 5 minutes and neonatal survival in neonates considered at-risk. This association is strong, even when stratifying the analyses across gestational age categories. These findings highlight the usefulness of the Apgar score at 5 minutes for decision-making processes for health professionals during the early care of at-risk neonates.

## What is already known?

⍰ Apgar score is associated with the risk of neonatal and infant death.
⍰ Apgar score helps to add efficacy of immediate resuscitative measures.

## What this study adds?

⍰ There is a strong direct association between Apgar score at 5 minutes and neonatal survival in neonates considered at-risk.
⍰ These findings highlight the usefulness of the Apgar score at 5 minutes for decision-making processes for health professionals during the early care of at-risk neonates.

## Data Availability

The Score Bebe project grants access to the main study results and data by making a request through the correspondence author.

## Footnotes

### Contributorship statement

IDE takes responsibility for (is the guarantor of) the content of the manuscript, including the data and analyses. IDE led the study, performed the data cleaning, the statistical analysis, and drafted the manuscript. All authors contributed to *(i)* the conception, hypotheses delineation, design of the study, acquisition of the data or analysis and interpretation of results, *(ii)* critically revised the article, *(iii)* approved the final version to be published, and *(iv)* agreed to be accountable for all aspects of the work.

### Patient consent for publication

Not required.

## Acknowledgments

Authors thank the input from the “Score Bebé” project members. We also acknowledge the contributions from the Ministry of Public Health of Ecuador (*Ministerio de Salud Pública del Ecuador, MSP*).

## Data sharing statement

The “Score Bebé” project grants access to the main study results and data by making a request through the correspondence author.

## References

1. Apgar, V. A proposal for a new method of evaluation of the newborn infant. Anesth. Analg. 120, 1056–1059 (2015).

2. Nelson, K. B. & Ellenberg, J. H. Apgar scores as predictors of chronic neurologic disability. Pediatrics 68, 36–44 (1981).

3. Iliodromiti, S., MacKay, D. F., Smith, G. C. S., Pell, J. P. & Nelson, S. M. Apgar score and the risk of cause-specific infant mortality: A population-based cohort study. Lancet 384, 1749–1755 (2014).

4. Bovbjerg, M. L., Dissanayake, M. V., Cheyney, M., Brown, J. & Snowden, J. M. Utility of the 5-Minute Apgar Score as a Research Endpoint. Am. J. Epidemiol. 188, 1695–1704 (2019).

5. Razaz, N. et al. Five-minute Apgar score as a marker for developmental vulnerability at 5 years of age. Arch. Dis. Child. Fetal Neonatal Ed. 101, F114–F120 (2016).

6. Phalen, A. G., Kirkby, S. & Dysart, K. The 5-Minute Apgar Score. J. Perinat. Neonatal Nurs. 26, 166–171 (2012).

7. The American College of Obstetricians and Gynaecologists. Executive summary: Neonatal Encephalopathy and Neurologic Outcome, Second Edition. Obstet. Gynecol. 123, 896–901 (2014).

8. Cnattingius, S., Johansson, S. & Razaz, N. Apgar Score and Risk of Neonatal Death among Preterm Infants. N. Engl. J. Med. 383, 49–57 (2020).

9. Li, F. et al. The Apgar Score and Infant Mortality. PLoS One 8, 1–8 (2013).

10. Bewick, V., Cheek, L. & Ball, J. Statistics review 12: Survival analysis. Crit. Care 8, 389–394 (2004).

11. Marchand, B., Tello, B., Dueñas-Espín, I. & Bonifaz, B. Atención integral a las enfermedades prevalentes de la infancia (AIEPI) clínico. Cuadros de procedimientos. (Ministerio de Salud Pública del Ecuador, 2018).

12. Prócel, M. G. et al. Manual de atención integral a la niñez. (Ministerio de Salud Pública del Ecuador, 2018).

13. American College of Obstetricians. ACOG Practice bulletin no. 134: fetal growth restriction. Obstet. Gynecol. 121, 1122–1133 (2013).

14. Villar, J. et al. Postnatal growth standards for preterm infants: the Preterm Postnatal Follow-up Study of the INTERGROWTH-21(st) Project. Lancet Glob Heal. 3, e681–91 (2015).

15. Mandel, M. Inverse Probability Weighted Cox Regression for Doubly Truncated Data. Biometrics 74, 481–487 (2018).

16. Tyson, J. E., Parikh, N. A., Langer, J., Green, C. & Higgins, R. D. Intensive care for extreme prematurity-moving beyond gestational age. Obstet. Gynecol. Surv. 63, 555–556 (2008).

