## Supplementary material for "Strong association between Apgar score at 5 minutes and neonatal survival among at-risk neonates": online supplementary

(ONLINE SUPPLEMENTARY MATERIAL)

Dueñas-Espín, I. *et al*.

### Process of building the database

The database was built based on the information collected from all hospitals in the Public Network of Health Establishments (made up of all public hospitals and its associates) and the Private Network of Health Establishments, including all Specialty Hospitals of Gynaecology and Obstetrics, as well as any hospital that attends births. The database is built from systematic reports from the health establishments whenever there is a newborn death (≤28 days of life). The reports are sent in through a pre-established form by using a unique username and password for each establishment. This username is assigned by the system manager (DNVE).

#### Actors of the health system which are obligated to report neonatal deaths

All public and private health establishments in the country are obligated to report any neonatal death in the first 24 hours after the event.

#### Established criteria for reporting neonatal deaths

There are two pre-established forms and two ways to notify neonatal deaths in the system. The first one is a simplified report where all neonatal deaths are reported. The second one registers both the information from the simplified report and other additional information, including criteria that show preventability. This data includes some anonymized information from the mother.

#### Verifying information

There is an epidemiologist responsible for each political-administrative zone. This epidemiologist has a username and password that gives him/her access to all the information reported in the designated area. The epidemiologist is responsible for reviewing the information and detecting incongruences, by verifying with those who report it.

### Figure 1S.- Diagram which shows the number of neonates registered, excluded, and included in the analysis.

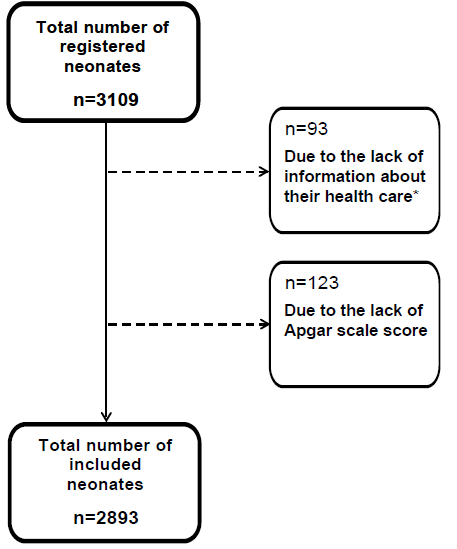

### Table 1S. – Individual and contextual variables of the study population.

| **Variables** | **Whole included population**  **N=2893** |
| --- | --- |
| **Individual variables^a^** |  |
| Female sex, n (%)**^b^** | 1380 (48) |
| Gestational age in weeks, median (P25 to P75) **^b^** | 31 (27 to 36) |
| *≤27 wk*, n (%) | 742 (26) |
| *>27 to <31 wk.* , n (%) | 709 (25) |
| *>31 to 36 wk*, n (%)*.* | 751 (26) |
| *>36 to 38 wk.* , n (%) | 364 (13) |
| *>38 to <41 wk.* , n (%) | 364 (13) |
| *>41 wk.* , n (%) | 66 (2) |
| Birth weight in g, mean (SD) **^b^** | 1651 (922) |
| *<750 g, n (%)* | 445 (15) |
| *750 to <1000 g, n (%)* | 477 (17) |
| *1000 to <1500 g, n (%)* | 644 (22) |
| *1500 to <2500 g, n (%)* | 684 (24) |
| *2500 to <4000 g, n (%)* | 607 (21) |
| *≥4000 g, n (%)* | 24 (1) |
| Small for gestational age, n (%)**^b^** | 759 (28) |
| Type of delivery **^b^** |  |
| *C-section, n (%)* | 1679 (59) |
| *Vaginal delivery, n (%)* | 1023 (36) |
| *Dystocic delivery, n (%)* | 159 (6) |
| Apgar scale score at 5 minutes, p50 (P25 to P75) | 6 (4 to 8) |
| *9 to 10, n (%)* | 547 (19) |
| *7 to 9, n (%)* | 864 (30) |
| *5 to 6, n (%)* | 719 (25) |
| *3 to 4, n (%)* | 333 (12) |
| *0 to 2, n (%)* | 430 (15) |
| Comorbidities |  |
| *Asphyxia related disorders, n (%)* | 695 (24) |
| *Malformations, n (%)* | 678 (23) |
| *Prematurity related disorders, n (%)* | 883 (31) |
| *Infectious diseases, n (%)* | 533 (18) |
| *Other non-previously classified, n (%)* | 104 (4) |
| **Contextual variables^a^** |  |
| Type of health care center |  |
| *Private medical care, n (%)* | 773 (27) |
| *Public medical care, n (%)* | 2120 (73) |
| Level of care**^b^** |  |
| *Primary care unit, n (%)* | 52 (2) |
| *Secondary care unit, n (%)* | 1271 (44) |
| *Tertiary care unit, n (%)* | 1557 (54) |
| * | |

### Figure 2S.- Histogram of the distribution of neonatal deaths per Apgar scale score at 5 minutes.

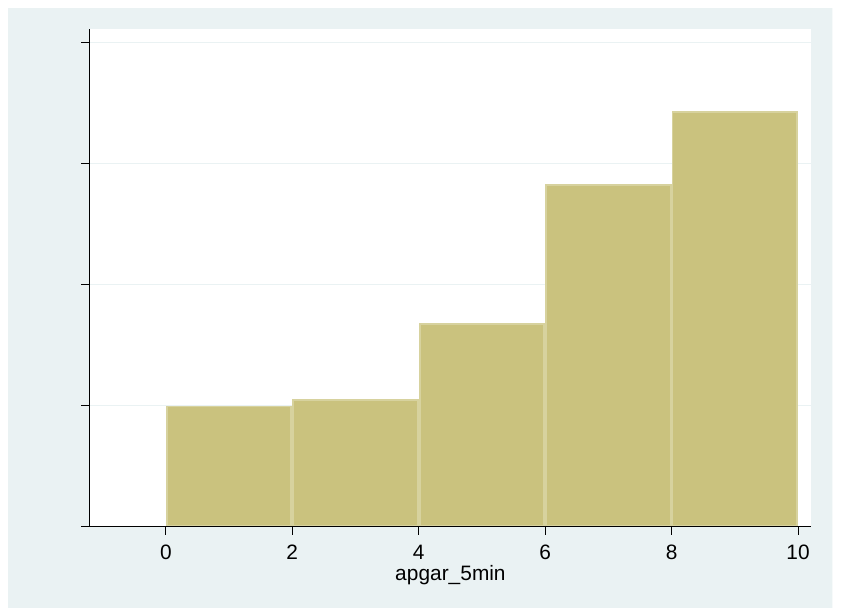

Figure 3S .- *Kaplan Meier survival estimates by Apgar at 5 minutes score, stratified by gestational age categories.*

| 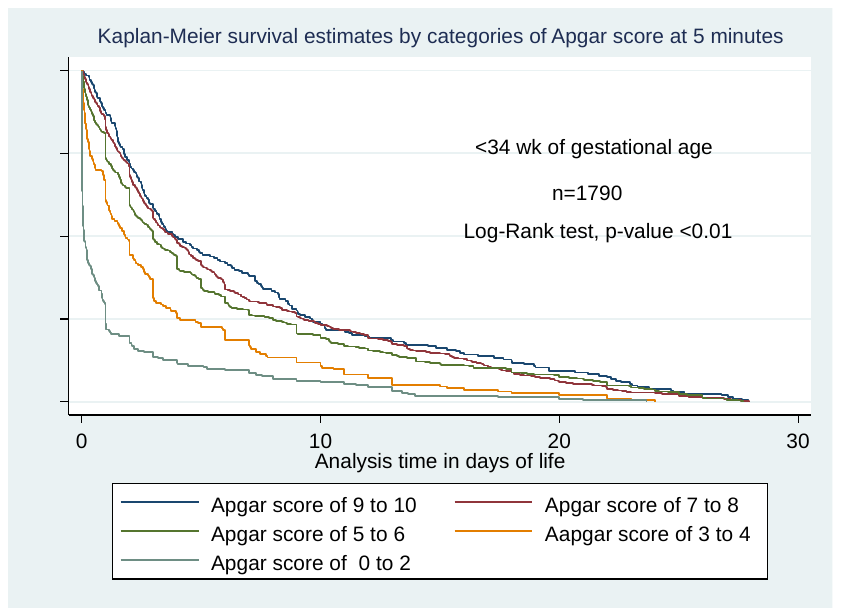 | 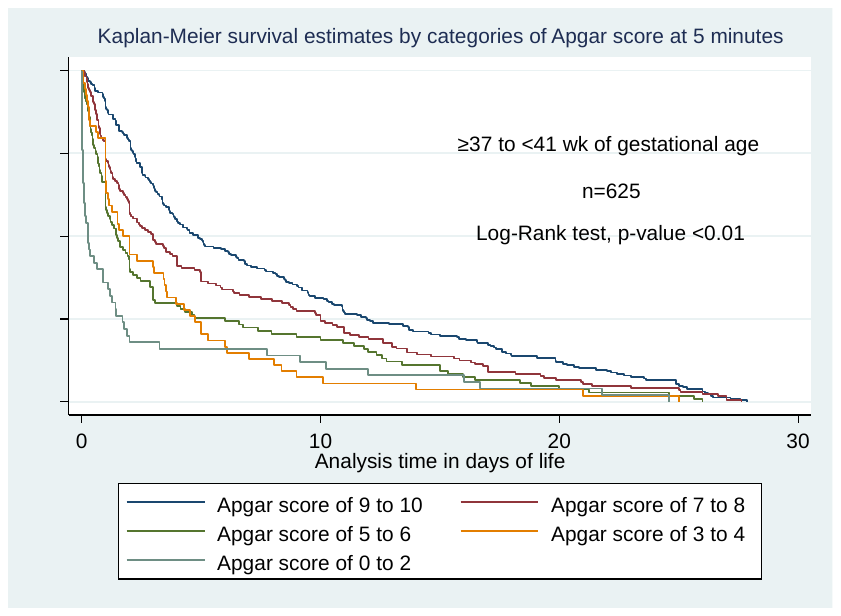 |
| --- | --- |
| 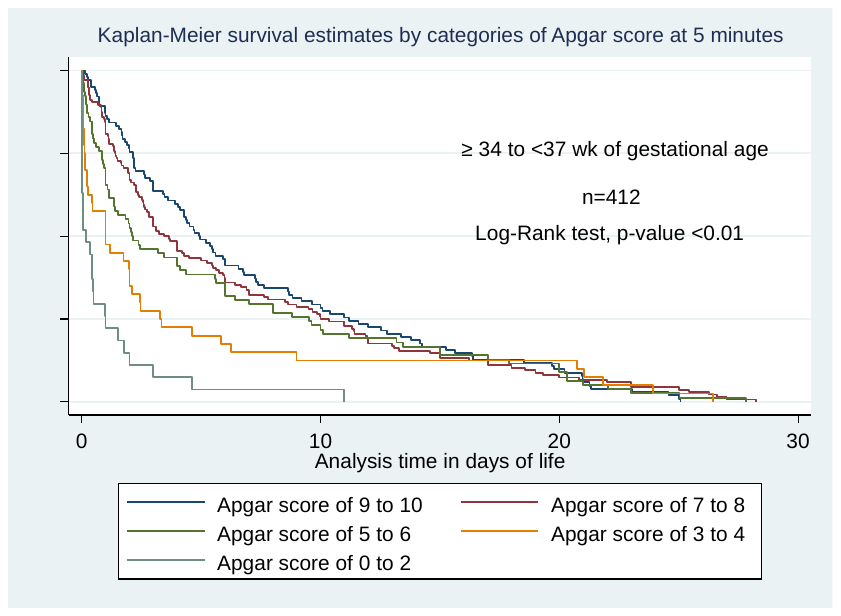 | 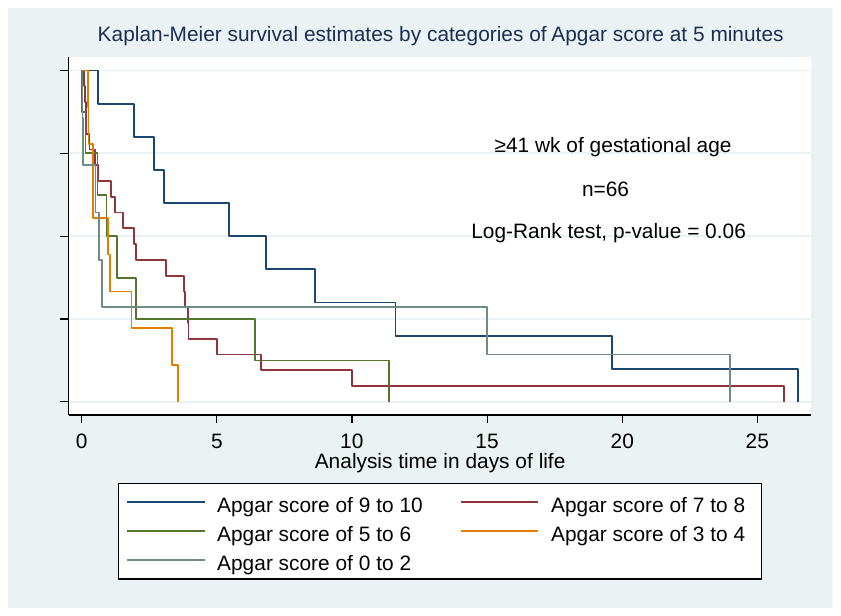 |

### Figure 4S.- Kaplan Meier survival estimates by Apgar at 5 minutes score categories, stratified by gestational age categories and after artificial cutting-off the follow up at 21 days

| 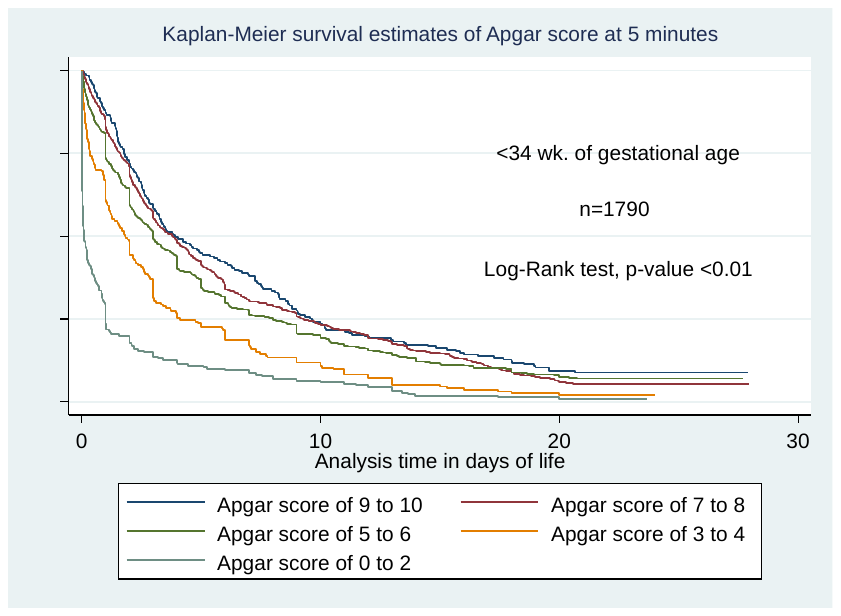 | 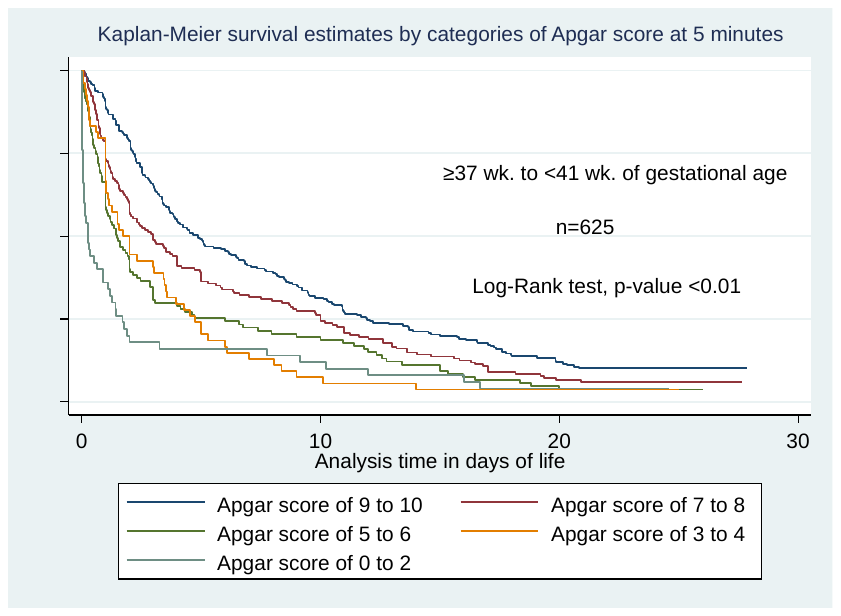 |
| --- | --- |
| 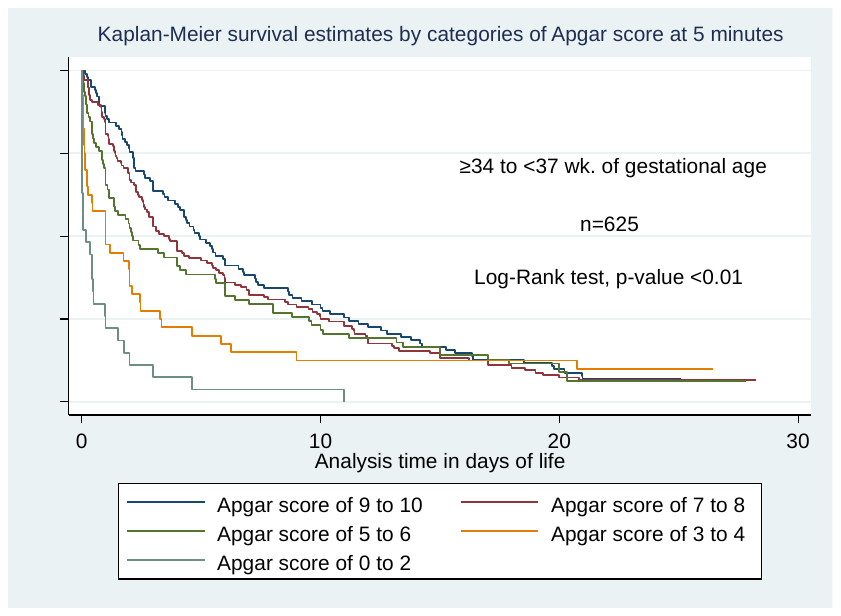 | 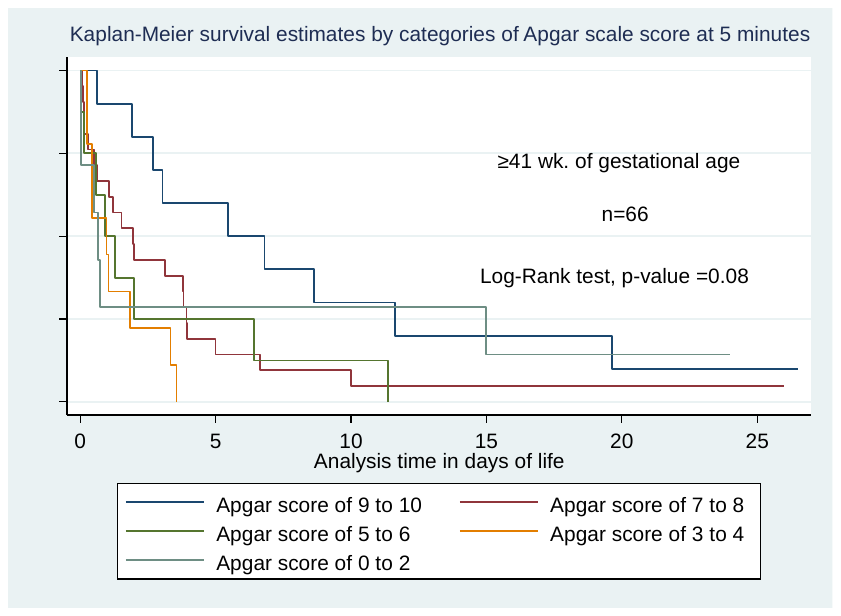 |

### Table 2S.- Adjusted neonatal mortality adjusted hazard ratios per each category of Apgar scale score at 5 minutes, according to fixed-effects multivariate Cox proportional hazards models, excluging (i) asphyxia related disorders, (ii) congenital malformations, (iii) prematurity related disorders, (iv) infectious disorders, and (v) other disorders non previously classified.

|  | **Adjusted Hazard Ratios^a^** | | | | | | | | | |
| --- | --- | --- | --- | --- | --- | --- | --- | --- | --- | --- |
| **Apgar scale score at 5 minutes** | **Asphyxia related disorders** | ***p-value*** | **Congenital malformations** | ***p-value*** | **Prematurity related disorders** | ***p-value*** | **Infectious disorders** | ***p-value*** | **Other disorders non previously classified** | ***p-value*** |
| *9 to 10, (ref.)* | 1 | - | 1 | - | 1 | - | 1 | - | 1 | - |
| *7 to 9* | 1.31 (1.15 to 1.50) | <0.01 | 1.09 (0.95 to 1.25) | 0.22 | 1.24 (1.08 to 1.43) | <0.01 | 1.34 (1.17 to 1.54) | <0.01 | 1.31 (1.16 to 1.48) | <0.01 |
| *5 to 6* | 1.57 (1.35 to 1.83) | <0.01 | 1.27 (1.09 to 1.49) | <0.01 | 1.43 (1.22 to 1.66) | <0.01 | 1.62 (1.39 to 1.88) | <0.01 | 1.51 (1.32 to 1.73) | <0.01 |
| *3 to 4* | 2.48 (2.04 to 3.01) | <0.01 | 1.64 (1.36 to 1.97) | <0.01 | 1.81 (1.50 to 2.18) | <0.01 | 2.14 (1.79 to 2.56) | <0.01 | 2.05 (1.74 to 2.41) | <0.01 |
| *0 to 2* | 4.53 (3.75 to 5.46) | <0.01 | 2.65 (2.21 to 3.18) | <0.01 | 2.93 (2.41 to 3.56) | <0.01 | 3.62 (3.04 to 4.30) | <0.01 | 3.33 (2.83 to 3.91) | <0.01 |
| *p-for-trend* | 1.42 (1.36 to 1.49) | <0.01 | 1.26 (1.21 to 1.32) | <0.01 | 1.27 (1.22 to 1.33) | <0.01 | 1.35 (1.30 to 1.40) | <0.01 | 1.32 (1.27 to 1.37) | <0.01 |
| **^a^** Each column is a different model adjusted by variables of Table 2: individual variables: *(i)* gestational age, *(ii)* birth weight, small for gestational age, *(iii)* type of delivery, *(iv)* comorbidities; and contextual variables: *(i)* type of health care center , *(ii)* public or private care, *(iii)* level of care. Please see main text for details. | | | | | | | | | | |
